# Exome sequencing as a first-tier test for copy number variant detection : retrospective evaluation and prospective screening in 2418 cases

**DOI:** 10.1101/2021.10.14.21264732

**Authors:** Quentin Testard, Xavier Vanhoye, Kevin Yauy, Marie-Emmanuelle Naud, Gaelle Vieville, Francis Rousseau, Benjamin Dauriat, Valentine Marquet, Sylvie Bourthoumieu, David Genevieve, Vincent Gatinois, Constance Wells, Marjolaine Willems, Christine Coubes, Lucile Pinson, Rodolphe Dard, Aude Tessier, Bérénice Hervé, François Vialard, Ines Harzallah, Renaud Touraine, Benjamin Cogné, Wallid Deb, Thomas Besnard, OIivier Pichon, Béatrice Laudier, Laurent Mesnard, Alice Doreille, Tiffany Busa, Chantal Missirian, Véronique Satre, Charles Coutton, Tristan Celse, Radu Harbuz, Laure Raymond, Jean-François Taly, Julien Thevenon

## Abstract

**Purpose:** Despite exome (ES) or genome sequencing (GS) availability, chromosomal microarray (CMA) remains the first-line diagnostic tests in most rare disorders diagnostic work-up, looking for Copy-number variations (CNV), with a diagnostic yield of 10-20%. The question of the equivalence of CMA and ES in CNV calling is an organisational and economic question, especially when ordering a GS after a negative CMA and/or ES.

**Methods:** This work measures the equivalence between CMA and GATK4 exome sequencing depth of coverage method in detecting coding CNV on a retrospective cohort of 615 unrelated individuals. A prospective detection of ES CNV on a cohort of 2418 unrelated individuals, including the 615 individuals from the validation cohort, was performed.

**Results:** On the retrospective validation cohort every CNV was accurately detected (64/64 events). In the prospective cohort, 32 diagnostics were performed among the 2418 individuals with CNVs ranging from 704bp to aneuploidy. An incidental finding was reported. The overall increase in diagnostic yield was of 1.7%, varying from 1.2% in individuals with multiple congenital anomalies to 1.9% in individuals with chronic kidney failure.

**Conclusions:** Combining SNV and CNV detection increases the suitability of exome sequencing as a first-tier diagnostic test for suspected rare mendelian disorders. Before considering the prescription of a GS after a negative ES, a careful reanalysis with updated CNV calling and SNV annotation should be considered.

## INTRODUCTION

Copy Number Variants (CNV) represent the imbalance of the genomic material compared to the reference genome, resulting in an increase or decrease in genomic material. CNVs vary in size, although they are defined as variants with a minimum size of 1 kb^1^. Adoption of Chromosomal Microarray Analysis^2^ (CMA) techniques have proven invaluable in discovering pathogenic CNVs in a wide variety of diseases, especially for diagnosing multiple congenital anomalies (MCA) or Intellectual Disability Disorder (IDD). In routine practice, a diagnostic yield of ∼15% is reached for patients with intellectual disability disorder or MCA, and can be attributed to large CNVs (> 100 kb)^3^. Despite the rapid adoption of next generation sequencing, standard chromosomal analysis and CMA remain the first-tier tests for most rare disorders diagnostic work-up ^4,5^.

In practice, the average resolution of CMA technologies implemented in laboratories is about 50 kb^3^. In theory, Genome Sequencing (GS) CNV calling is the golden path for CNV calling. However, exome sequencing is notably widespread and more affordable, thus an accurate CNV calling should be advised on existing data before ordering an additional diagnostic test.

Although ES has intrinsic limitations, common problems are shared by GS and ES in calling CNV such as extreme GC contents or low complexity regions. In GS, algorithms strategies of type Depth of Coverage (DoC), Split Read, Discordant Pairs and Assembly^6^ can be used, whereas ES CNV calling tools can only use DoC. ES specifically encounters additional limitations regarding the targeted enrichment (known as capture bias), leading to non-uniform read depths impacting the reproducibility and robustness of CNV calling tools^7^. The ratio of read count between a test and a reference is usually preferred to a single-sample analysis, which could lead to many false positive^8^.

Numerous tools such as XHMM^9^, CODEX^10^, CANOES^11^, CoNIFER^12^ or ExomeDepth^8^ were developed when germline ES started to be democratized, none of them has really imposed itself as the reference tool. In January 2018 the Broad Institute released the fourth version of its GATK^13^ tool (GATK4) including several tools forming a CNV detection module^14^. This module is based on the principle of constructing a learning model from a cohort of patients DoC data that can be further reused.

This study presents an analytical validation framework for a clinical routine of GATK4 gCNV calling on ES data supported by a retrospective benchmark on 615 unrelated index cases with previously acquired CNVs. Results include the prospective screening for CNV in 1803 additional unrelated individuals with no previous CMA, totalizing 2418 individuals.

## PATIENTS AND METHODS

### Individuals gathering

Patients were ascertained in the diagnostic routine of the Eurofins Biomnis Laboratory (Lyon, France). The referring clinical centers included Nantes, Lyon, Montpellier, Paris (Tenon), Grenoble, Besançon, Saint Etienne, Limoges, Poissy, Marseille, Orléans and international laboratories (details provided in *Supplementary Material Table 1*). Patients provided written consent. A total of 2418 individuals were included in the work. Overall, 615 had CMA, MLPA or NGS-based data available as tabulated files and were used as the analytical retrospective validation cohort. Files formats were normalized during this study. For the remaining 1803 individuals, no question was asked regarding previously available CMA results, and are further referred as the prospective screening cohort in combination with the previous prospective cohort samples.

### ES capture sequencing

For all the 2418 probands, ES libraries were generated using standard procedures (*Supplementary Materials*) for 3 different capture protocols for sequencing Roche Medexome kit (n= 447), Twist Bioscience Human Comprehensive Exome kit + RefSeq + UTR spike (n= 988), Twist Bioscience Human Comprehensive Exome kit + RefSeq spike (n= 983). Libraries were sequenced on Illumina NextSeq 500 sequencers in paired-end mode (2 × 76bp).

### ES analysis for CNV calling

Exome Sequencing data was mapped against the hg38 genome, following the Broad Institute GATK best practice guidelines ^15^. CNV calls were performed with the GATK4 CNV calling module. Fine-tuning of ES learning model creation was performed according to parameters provided by the Broad Institute teams (shown in Supplementary Material). It was therefore decided to divide the calling target into 4 bins with the GATK IntervalListTools in order to run four instances of the GermlineCNVCaller in parallel on our computing infrastructure. The full methodology of model building is available in the *Supplementary Material*. Each VCFs were then annotated with AnnotSV^16^ version 2.5.1 to add crucial metadata for interpretation by the clinician. The output files by AnnotSV were processed by an in-house Python script to keep only the annotations of interest, but also to add the occurrence cohort counts of each CNV.

The diagnostic target represented 41 935 379 bp, defined by the merging of UCSC RefSeq and RefSeq Curated^17^ intervals, with 5’-3’ padding of 20bp. This diagnostic target included 21450 genes with 198188 exonic intervals.

For all samples, CNV were analyzed at the same time as SNV analysis. SNV interpretation was done following ACMG recommendations^18^. CNVs were prioritized based on their frequency in our cohort, and in DGV^19^; the inclusion of an OMIM Morbid gene; the quality metrics of the CNV and the inheritance of the CNV. Recurrent CNV were specifically analyzed according to gene content and recurrent CNV list of the French AChroPuce consortium (https://acpa-achropuce.com/).

### Analytical retrospective validation cohort

Biological results from 615 individuals with previously identified clinically relevant CNV were gathered and compared to CNV detection by ES. To ensure comparable results across detection techniques, only coding CNV were compared. Overall, 72 CNVs were considered as clinically relevant. 64 CNV were used for comparison, either classified as VUS, likely pathogenic or pathogenic. Frequent polymorphisms and technical artefacts may be confusing and were excluded from the analysis. The 64 CNVs included 30 loss, 31 gain (including a XXY phenotype) and 3 VUS with a copy number of 2 (chromosome X), with sizes ranging from an intragenic single exon deletion to large anomalies including aneuploidy (summarized in *Supplementary material table 2*).

### Prospective screening cohort

Prospective cohort included 2418 individuals. CNVs were called only on ES data. Each CNV larger than 1 Mb was individually interpreted. Regarding smaller CNV, filtering was performed (i) on the quality score QA > 20 and QS > 20; (ii) overlapping or impacting a gene referenced in the OMIM database with suspected or demonstrated dosage sensitivity (pLI >0.9); (iii) autosomal dominant inheritance for heterozygous CNV inheritance. Every homozygous and hemizygous CNV were considered. Each filtered CNV was interpreted and classified. Downstream CNV validations were performed by the referring centers using standard procedures.

## RESULTS

### Statistical description of CNV calls

Across capture kits, the distribution of the CNVs larger than 50kb number was varying from an average of 5-10 events. The median number of CNVs smaller than 50kb varied from 31-36 across capture kits (*Figure 1A*). The median number of CNV encompassing an OMIM morbid gene was comparable across capture kits. For morbid CNVs, their distribution is comparable between the 3 models, with a median of 4 (< 50kb) or 1 (> 50kb) (*Figure 1B*).

**Figure 1.**
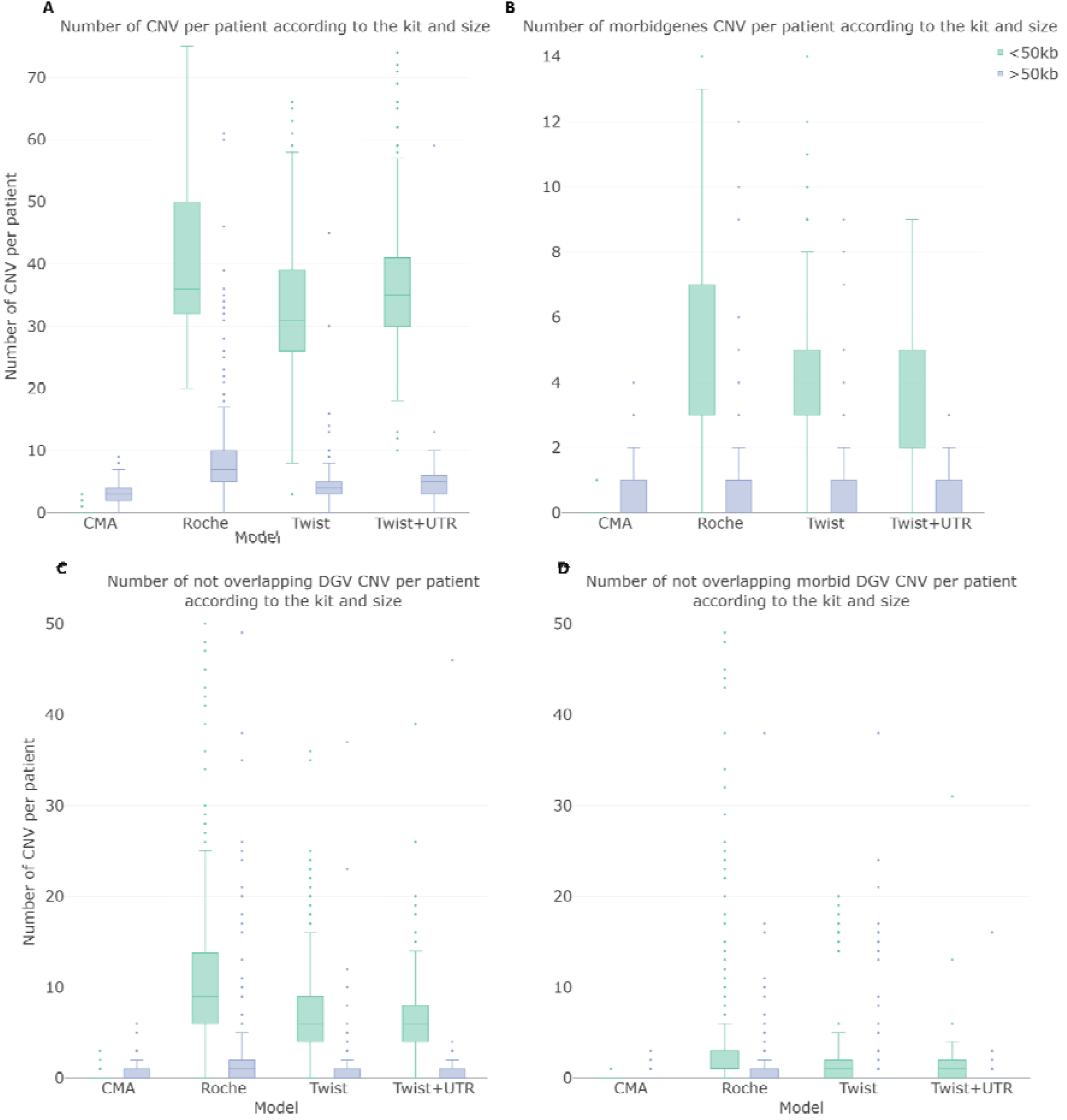
Distribution of the number of called CNV. (A) The total number of CNV, (B) the number of CNV containing at least one morbid gene, (C) the number of CNV not present in DGV, (D) the number of CNV containing at least one morbid gene not present in DGV, per patient according to the CNV size and the model used compared to CMA data. CMA (n=300), Roche (n=511), Twist (n=1154), Twist UTR (n=383).

Finally, detected CNVs were intersected with the DGV database. Intervals were considered comparable when at least 80% of reciprocal overlap was observed. A median of 75,56%, 77,78% and 75,76% (Roche, Twist, Twist+UTR) of detected CNVs were referenced in the DGV database (*Figure 1C*). A median of one CNV overlapping an OMIM morbid gene and absent from the DGV database was observed (*Figure 1D*).

### Defining the model size for ES-CNV calling

From the Twist model data set (n = 1154), several models were built of different sizes and random data (50, 100, 150, 200, 300, 600 samples), with three subsamples for each size condition. Then, from the Twist data set, 154 samples were randomly selected and were used as a fixed cohort. Iteratively, CNVs were called on those samples against the previously constructed models (*Figure 2*).

**Figure 2.**
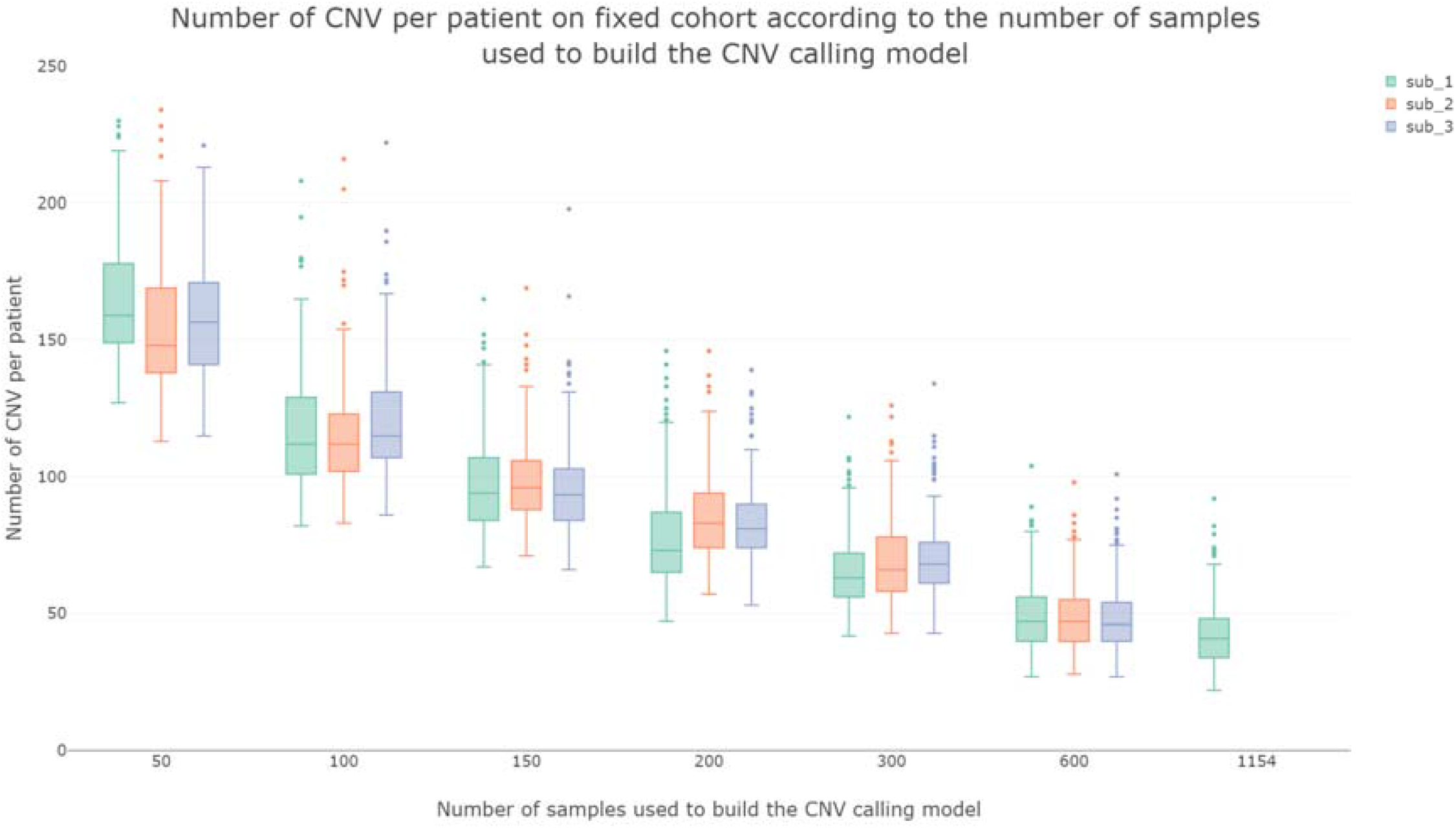
Distribution of the number of CNVs per patient in the cohort of 154 fixed patients according to model size and subsampling. 3 sub-samples (sub 1-3) of built CNV calling models consisting of 50 to 600 samples sequenced with the Twist Human Core Exome kit. CNV reused to call CNV 154 randomly selected samples (the same samples for every model) compared to the results of the Twist model consisting of 1154 samples on these 154 samples.

The lower the number of samples used to build the model, the higher the average number of CNVs per patient and vice versa (*Figure 2*). In addition, the smaller the models, the more variable are the distributions between the subsamples. Among the 1154 samples, and independently from the calling model, 23 individuals were continuously leading to high numbers of CNV calls (> 200).

### Isolating outliers of the ES-CNV calling pipeline

Among the whole cohort (2418 samples), 2275 individuals had fewer than 200 events. 143 samples were leading to an excess of CNV calls across capture kits and calling models. The distribution of CNV counts is represented by *Supplementary material Figure 4*. These 143 outlier samples were excluded from the interpretation and further analysis. Among the 143 samples, 66 were concentrated in seven sequencing runs with technical issues; 67 samples were DNA received from collaborators (60 DNA extracted from blood and 7 DNA extracted from tissues); 10 were blood samples received by the laboratory.

### Defining recurrent uncallable regions

ES CNV calling was unable to quantify the copy number ratio for a significant portion of the diagnostic target, 10 and 11% for Twist capture kits and 8.76% for the Roche kit. Focusing on the 3593 genes of the OMIM morbidmap identified 32 genes totally uncallable for coding CNV (*AHDC1, AMER1, BBS12, CHAMP1, CRYAA, CSF2RA, DOLK, FLRT3, FZD2, GP1BA, HPS6, IRF2BPL, IRS4, KCNA1, KCNA4, KCNA5, MAGEL2, MKRN3, MYORG, PIGW, POMGNT2, RAG2, SAMD9, SAMD9L, SLC18A3, SLITRK1, SLITRK6, THBD, TRIM32, UBQLN2, ZNF469, MARCH2*). Across capture kits and each calling model, an average of 410 genes are partially represented and CNV calling might be impacted (*Supplementary material Figure 5*).

### Analytical retrospective validation cohort

Overall, 615 samples were available. Twenty-five (4.0%) samples were excluded from the analysis because they were classified as outliers. Among the 72 selected CNVs, 8 were excluded because they were localised in intergenic regions or in a previously defined uncallable region (*Supplementary material Figure 6*). For the 590 remaining samples, the 64 CNVs were accurately detected and genotyped (*Supplementary material table 2*). No additional large and rare CNV was reported.

### Prospective screening cohort

Among the 2418 individuals, 30 CNV and 2 aneuploidies were diagnosed. Among the 1787 individuals with MCA/IDD, 20 diagnoses were performed. Among the 631 individuals with chronic kidney failure, 12 diagnoses were performed (*Supplementary material table 3, 4*). Regarding the 22 pathogenic or likely pathogenic CNV larger than 50 kB, ES was the first genetic investigation.

Patient 4 was presenting with chronic kidney failure and kidney cysts in adulthood, revealed an intragenic deletion of *COL4A3* at heterozygous state. BAM viewing emphasized breakpoints in exon 9 (*Figure 3*). Breakpoints were verified using Sanger sequencing, allowing characterization of the variation : NC_000002.12:g.227248049_227251231del; NM_000091.4:c.469-394_609+29del. Small pathogenic or likely pathogenic CNV in genes of recessive inheritance, associated with a pathogenic or likely pathogenic SNV on the other allele for 2 patients were detected. Patient 5, presenting with dilated cardiomyopathy and facial dysmorphism, carried NM_006663.3(*PPP1R13L*):c.1871_1872del; p.(Arg624Profs*119), maternally inherited, and intragenic duplication of *PPP1R13L* (duplication of exon 2 to exon 7, of 13). Patient 6 presented with growth delay, facial dysmorphia, delayed psychomotor development, hyperextensibility, cortical atrophy, thin corpus callosum and hypomyelination. ES detected a deletion of the whole *PYCR2* gene, maternally inherited and a hemizygous point variant paternally inherited : NM013328.3(*PYCR2*):c.751C>T; p.(Arg251Cys).

**Figure 3.**
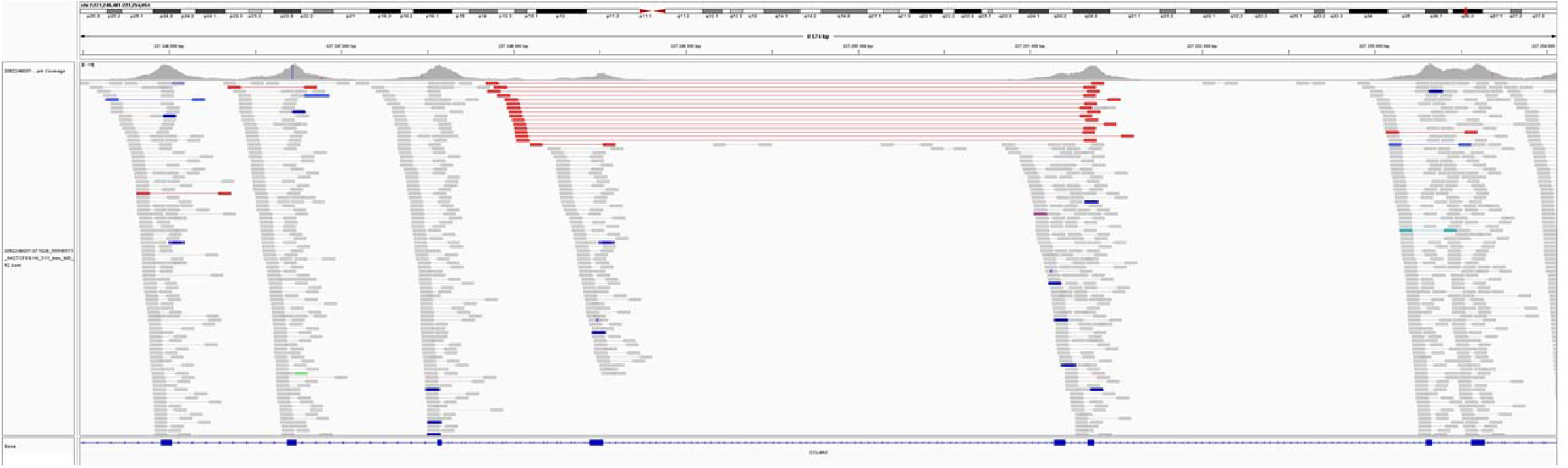
BAM visualisation of Intragenic heterozygous deletion of COL4A3 exon 9. Reads colored, oriented and sorted by insert size with IGV software.

## DISCUSSION

This study assessed the analytical validity of gCNV calling in an ES routine based on a 615 individuals retrospective validation cohort and demonstrated the positive impact on ES diagnostic yield through the screening of 2418 individuals. In this first-tier ES routine, CNV calling identified 2 aneuploidy, 22 large CNV and 8 small CNV.

The 64 CNV gathered from the retrospective validation cohort were accurately detected and genotyped by the ES procedure. Previous study had demonstrated the equivalence of ES against CMA ^20^. Another published cohort included 147 samples with 102 CNV, and they performed comparison between aCGH CNV detection and CANOES CNV detection^11^. The recall was 87.2% (89/102). They suggested that the missed CNV by ES might be secondary to the capture design or size of the event with only 1 or 2 targets ^11^. Our retrospective validation dataset included very small events such as hemizygous deletion of one exon in *DMD* gene or gain of one exon in *IL1RAPL1* (respectively for individuals 2 and 1, *Supplementary material table 2)*, which were accurately identified. These observations suggest that this work may add an important validation of the procedure for a clinical ES routine.

In the validation cohort, the exhaustive detection of CNV may be secondary to the preliminary definition of predictive limitations of the procedure. These limitations included the definition of uncallable regions, and the prediction of aberrant and noisy samples. This study did not aim at deciphering the underlying causes for these limitations.

Analysis of outliers of CNV-ES detection reveal that our workflow is robust and suitable for routine diagnosis, with 6% of failed samples (143/2418). Most of those outlier samples could be explained by pre-analytical or analytical issues. Only 10 blood samples (among 1698) were classified as “outliers”. This failure rate of 0.6% is acceptable and comparable or below those of CMA in our practice. To further investigate those outliers, we analyzed CNV calls for outliers of the validation cohort : all medically relevant CNV were properly called, with high quality metrics. Those data suggest that CNV calling is possible for samples initially classified as outliers, but require intensive filtration and interpretation, to distinguish authentic CNV and background noise.

Tools to model coverage distributions across exons are widespread in the clinical bioinformatics community. On the other hand, the possibility of being able to build a learning model, and then to reuse it later on, seems genuinely new. The performances of the CNV calling models are certainly correlated to the number of data items that were used to build them. However, two models built with the same number of data and different sequencing depths will have different results. It is therefore more likely that the efficiency of the model is correlated to the cumulative sequencing depth of the data that compose it as well as their homogeneity across individuals. With the current sequencing data generation processes in our lab, if we ever had to reconstruct a model, the number of samples required would most likely be around 300.

GS has been proven to be more efficient for diagnosis than ES, both for SNV and CNV^21–24^. Indeed, in addition to being able to detect exonic, intronic and intergenic SNVs and indels, GS can more accurately detect exonic, intronic and intergenic structural variants. Unlike ES, the production of GS data does not require prior amplification or capture steps. This limits the variability of depth between exons, and virtually extinct the uncallable regions. Nevertheless, even if the set of uncaptured zones represents about 4 mb or 10% compared to the defined medical target in this study. However, only 0.9% of morbid genes have their entire sequence in the blind areas of our pipeline. Copy number variations in these genes will not be detected. However, large CNVs encompassing such genes might be detected.

Careful examination of the data generated by the pipeline allowed identification of causing-disease CNV for 33 patients. One incidental finding among the sequenced probands was identified (see Supplementary Table 3).

Among these 32 positive results, 8 individuals had a negative CMA before ES prescription. In the neurodevelopmental disorder cohort, the added diagnosis range is 1,2% (20/1787). This percentage is relatively low compared with the yield of >10% reported for genomic microarrays. This can easily be explained by the fact that the vast majority of patients with a neurodevelopmental disorder were previously screened negative for CNV microarray analysis, resulting in a depletion of pathogenic CNVs in this patient group. Clinically relevant CNVs were observed only in patients who had previously been screened on a (low-resolution) microarray platform or in patients who did not receive microarray-based CNV profiling. This percentage is consistent with previous studies analyzing exome based CNV calling within IDD cohorts (1.3%^25^; 1.6%^26^). Among individuals with chronic kidney failure, the diagnosis yield reaches 1,9 % (12/631). Only few data highlights the implication of CNV in renal disease. Previous studies demonstrated an added diagnosis range of 3.6% (2 of 56 patients)^25^ with CNV detection.

Of note, using an exome-wide CNV detection pipeline raises new incidental findings. We identified a deletion of 6 exons of LDLR (responsible for familial hypercholesterolemia [OMIM:# 143890)] for a patient referred for neurodevelopmental disorders.

The commitment to make ES a frontline analysis is not new^27^. On one hand, it has already been shown that ES can be much more efficient than traditional methods in terms of diagnostic rates as well as cost-effectiveness^28^. On the other hand, ES has already shown its superiority against some routine genetic analyses such as gene panels and single gene testing^24,29^. The ability to bundle the detection of exonic SNVs, Indels and CNVs make the ES strategy an extremely competitive and efficient first-tier analysis. In this cohort, two diagnoses were performed by combining CNV and SNV calling (0.08%, 2/2418). This observation is consistent with data from a large study of 12000 individuals combining CMA and ES for the identification of 17 diagnoses (0.11%)^30^. Despite limitations, thousands of exomes will be produced in the coming years for the diagnosis of rare disorders. A careful and updated analysis will enhance the diagnostic yield of the tests and will participate in reducing the diagnostic odyssey of patients with undiagnosed disorders.

This study highlights the technical validity and the clinical utility of exome-based CNV screening. Incorporation of CNV analysis in exome sequencing data-analysis pipelines increases the diagnostic yield of exome sequencing by up to 1,9%. Of importance, this increase in diagnostic yield is obtained without any additional direct laboratory costs. Combining SNV and CNV detection increases the suitability of exome sequencing as a first-tier diagnostic test for many, if not most, suspected genetic disorders. Before considering the prescription of a GS after a negative ES, a careful reanalysis with updated CNV calling and SNV annotation should be considered.

## Data Availability

Raw genetics data are not available. All data produced in the present work are contained in the manuscript.

