## Supplementary Materials for "Exome sequencing as a first-tier test for copy number variant detection : retrospective evaluation and prospective screening in 2418 cases"

| Prescribing center | Number of patient reference data provided |
| --- | --- |
| Nantes | 349 |
| Montpellier | 79 |
| Saint-Étienne | 59 |
| Limoges | 55 |
| Poissy | 50 |
| Marseille | 11 |
| Besançon | 2 |
| Orléans | 2 |
| Tenon | 1 |
| International Laboratories | 7 |
| Total | 615 |

**Supplementary Table 1.** Origin of reference CMA data from the validation cohort.

**DNA preparation for ES sequencing**

The genomic DNA is extracted from blood samples using the QIAsymphony® DSP DNA Mini Kit on a QIAsymphony instrument following the recommendation of QiaGen.

**QC DNA and normalisation**

The concentration of the genomic DNA is established using Optical density at 260 nm with a Spectrophotometer (DropSense 16).

After normalisation, 50 ng of genomic DNA are engaged in a library preparation step. The generation of the captured pooled libraries are prepared using the Human Comprehensive Exome kit delivered by Twist Bioscience following their recommendation.

**ES capture libraries preparation**

It consists of a first step that combines an enzymatic fragmentation, repair ends and dA tailing. The dA-tailed DNA fragments are then ligated to universal adapters and purify on magnetic beads. These purified fragments are amplified by PCR (7 cycles) using Universal Dual Index Primers (Truseq compatible) and the KAPA Hifi HotStart enzyme. After cycling the amplicons are purified on magnetic beads. Each library sample is quantified using the Thermo Fisher Scientific Qubit dsDNA Broad Range Quantitation Assay on Qubit (Thermo fisher Scientific) and the size range is validated on a migration on a Fragment Analyzer (Agilent) using the NGS Fragment analysis kit.

**CNV model building and reusing**

**ES pre-CNV calling target processing**

The GATK4 tools from the gCNV module used in our ES CNV calling models creation are shown in *Supp Fig 1*. Model creation was done by launching scripts and command lines, some steps were parallelized with the GNU Parallel^32^ tool. Some steps must have been taken beforehand to obtain the final CNV calling model target from our medical reference target, RefSeq^17^, in Picard^33^ INTERVAL_LIST format. The PreprocessIntervals tool was applied with a padding parameter, expanding to a maximum of 250 base pair (interval expansion stopped when it was likely to overlap with another interval) each interval of RefSeq. Then, a depth of coverage counts of the expanded target intervals of all patient BAM files included in the cohort was performed with the CollectReadCounts tool. Meanwhile, the percentage in GC of the extended target was calculated with the AnnotateIntervals tool. The annotated and extended targets with the cohort count dataset were provided to the FilterInterval tool, which filtered out intervals with extreme GC content, but also over or under-captured intervals in the count dataset. The final filtered target was used to recalculate all the patient coverage data of the cohort.

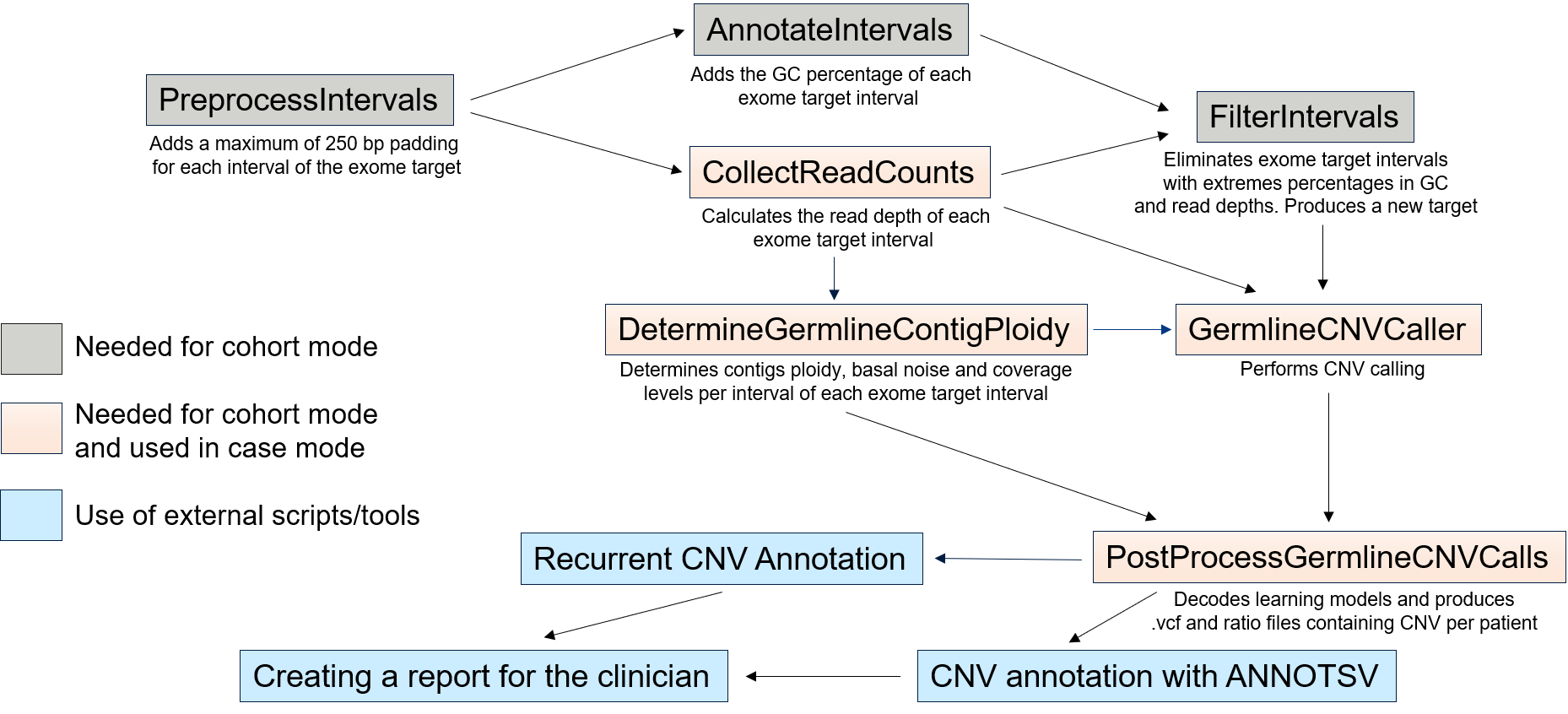

**Supplementary Figure 1.** gCNV calling workflow. Gray rectangles represent the preliminary steps in obtaining a suitable calling target for the pipeline, with areas which are not suitable for calling masked. Those steps are performed only once during the model creation. Orange rectangles represent the steps that create learning models in *COHORT* mode and reuse them in *CASE* mode. Yellow rectangles represent scripts or external tools to GATK4 used to add metadata to the produced *VCF* files. Only tools in the orange and blue rectangles are present in our production pipeline.

**ES learning model creation and CNV calling**

Patient count data and the filtered calling target were given in input to the DetermineGermline- ContigPloidy tool in *COHORT* mode. A DetermineGermlineContigPloidy learning model and call files were created from the cohort. Call files summarized the ploidy of every chromosome for each of the cohort patient. *COHORT* mode produced models and calls whereas *CASE* mode only produced calls by reusing a model created by a previous *COHORT* mode execution.

Patient count data, filtered calling target and DetermineGermlineContigPloidy model were provided as input to the GermlineCNVCaller tool. As above, it created a model from the cohort patient dataset and call files. The PostProcessGermlineCNVcalls tool converted the models and calls produced by the DetermineGermlineContigPloidy and GermlineCNVCaller into ratio files (with the estimated number of copies per interval for each patient), interval VCF files (reporting all intervals or bin from the filtered target), but also segment VCF files (merging contiguous intervals supposed to belong to the same original CNV) which were the files used afterwards.

**Learning model built and used for CNV calling**

Currently, 3 models are used in diagnostic routine, the Roche model built with data from the Roche protocol (n = 511), the Twist model built with data from the Twist Bioscience Human Core Exome kit + RefSeq spike protocol (n = 1154) and the Twist UTR model built with data from the Twist Bioscience Human Core Exome kit + RefSeq spike + UTR spike protocol (n = 383). Two additional models were used as a reference to study the performance of the models currently in production, but were not used for diagnosis. The Twist Legacy model, populated with the entire Twist protocol data set at the time of its creation (n = 432) and the Twist All model, containing the entire Twist and Twist UTR data set plus demultiplexing duplicates (n = 1730).

**CNV calling ES learning model re-use**

The reuse of the different machine learning models from the DetermineGermlineContigPloidy and GermlineCNVCaller tools was done within the same Nextflow pipeline that we used to produce the BAM files. Reusing models rather than building them was a much less demanding processor in terms of resources and computing time. The sequence of steps in the pipeline was the same as when the model was created, but these steps are performed within an automated production framework. The only difference between the two execution modes is that the CNV count of the VCF files produced in a model reuse is added to the CNV count of the cohort used to build the model, in order to have a more accurate recurrent CNV count.

**CNV Fine tuning model creation parameters before CNV calling results**

During our various preliminary tests with the gCNV calling pipeline of GATK4, we realized that the CNV calling results could be significantly improved, depending on the data, by changing some of the default parameters. We discussed with the Broad Institute team in charge of the pipeline development and we determined a list of parameters allegedly "optimized" for our exome sequencing data. We tested the impact of these parameters against the default parameters with the datasets we had at the time, *i.e.,* a Roche model consisting of 511 samples (still the model used for the Roche data, as we have not produced any new data using this wet lab protocol since then) and a Twist Legacy model consisting of 432 samples (samples that are fully included in the Twist model with 1154 samples constructed *a posteriori* with all the data produced with this wet lab protocol to date), results are shown in *Supp Fig 2*.

**Supplementary Figure 2.** Number of CNV in VCF file per patient (only CNV genotype different to 0) according to the set of parameters and the model used for the model construction (Roche n=511, Twist_Legacy n=432).
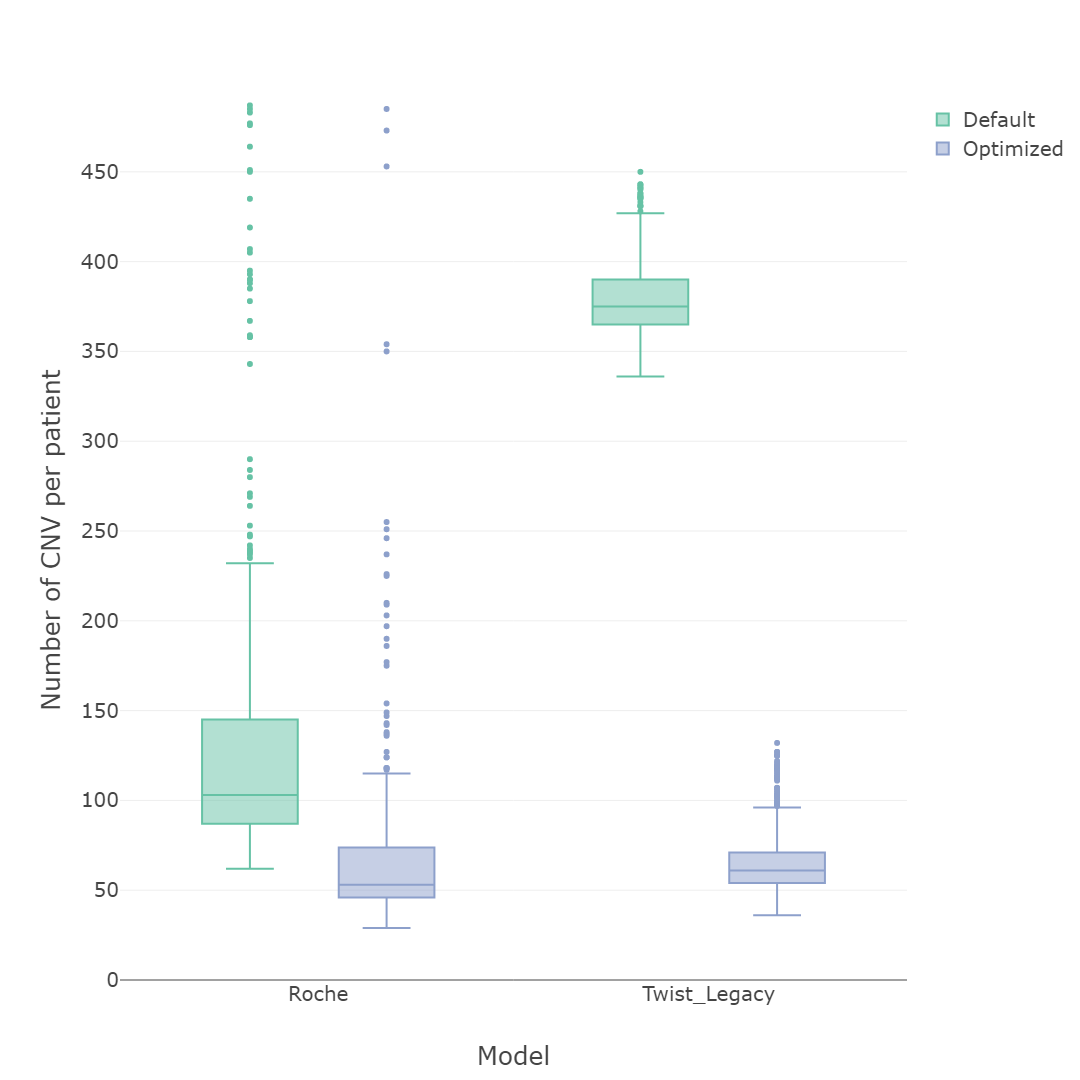

For both kits optimized parameters have a lower number of CNVs distribution per patient in regards to default parameters, which is expected for a more effective learning model and thus a more effective CNV calling model. Nevertheless, the mean or median number of CNVs per patient for the default condition is very different between the two kits, whereas using the optimized settings brings this number to about the same level. It is difficult to comment on CNV calling model differences, as the two datasets were produced with a different wet lab protocol, so they cannot be compared.

We have nevertheless tried to determine what could be the cause of these differences in profile. We first thought of an effect of patient recruitment bias that would lead to over-recruitment of poor-quality samples with too many CNVs. However, it is the Roche dataset that presents the most data with an atypical number of CNVs per patient, as it is the first exome data that we have produced ES data and on which we have learned the most. This does not correspond with a very high number of CNVs for the default condition of the Twist Legacy model compared to the Roche model. We compared many metrics between these two datasets and finally found that the real difference would be in the sequencing depth.

**Optimized parameters for DetermingermlineContigPloidy tool**

--sample-psi-scale 0.001
--global-psi-scale 0.05
--mean-bias-standard-deviation 1.0

**Optimized parameters for GermlineCNVCaller**

-adamax-beta-2 0.97
--caller-update-convergence-threshold 1e-06
--class-coherence-length 100000.0
--cnv-coherence-length 100000.0
--convergence-snr-trigger-threshold 0.2
--copy-number-posterior-expectation-mode EXACT
--depth-correction-tau 100.0
--interval-psi-scale 0.002
--learning-rate 0.03
--log-emission-sampling-median-rel-error 0.001
--log-emission-sampling-rounds 20
--log-mean-bias-standard-deviation 10.0
--max-advi-iter-first-epoch 5000
--max-advi-iter-subsequent-epochs 200
--max-bias-factors 16
--max-calling-iters 20
--max-training-epochs 20
 --min-training-epochs 5
--num-thermal-advi-iters 4000
--p-active 0.01
--p-alt 0.001
--sample-psi-scale 1e-07

**Effect of binned target on CNV calling results**

The creation of the learning model and more particularly the model of the GermlineCNVCaller step was a particularly long step and consumed computational resources, it was decided to divide the calling target to be able to parallelize their creation. We therefore divided our calling target into 4, 8 and 23 bins. For the first two conditions, we used GATK's IntervalListTools, which divides files in INTERVAL_LIST format into roughly equal parts. For the 23 bins condition, we made one bin per autosomal chromosome and one bin for the two gonosomal chromosomes.

We therefore divided our calling target into 4, 8 and 23 bins. For the first two conditions, we used GATK's IntervalListTools, which divides files in INTERVAL_LIST format into roughly equal parts. For the 23 bins condition, we made one bin per autosomal chromosome and one bin for the two gonosomal chromosomes.

To determine whether the number of bins had any effect on the outcome of CNV calling, we decided to recreate models by reusing the data from the Roche (n = 511) and Twist Legacy (n = 432) models and the different binned targets previously created. The results of the different models are shown in *Supp Fig 3*.

A slight decrease in the number of unexplained CNVs per patient is visible for the Twist Legacy, default, “8 bin”, condition. A clear decrease in the number of CNV per patient is visible for the condition Twist Legacy, default, “23 bin”, due to the fact that the chr2 bin could never be completed with GATK version 4.1.4.1 due to a bug known by the developers. No significant difference in distribution attributable to the number of bins is visible.

We have chosen for reasons of practicality related to the size of our computing infrastructure to create our models by dividing them into 4 bins.

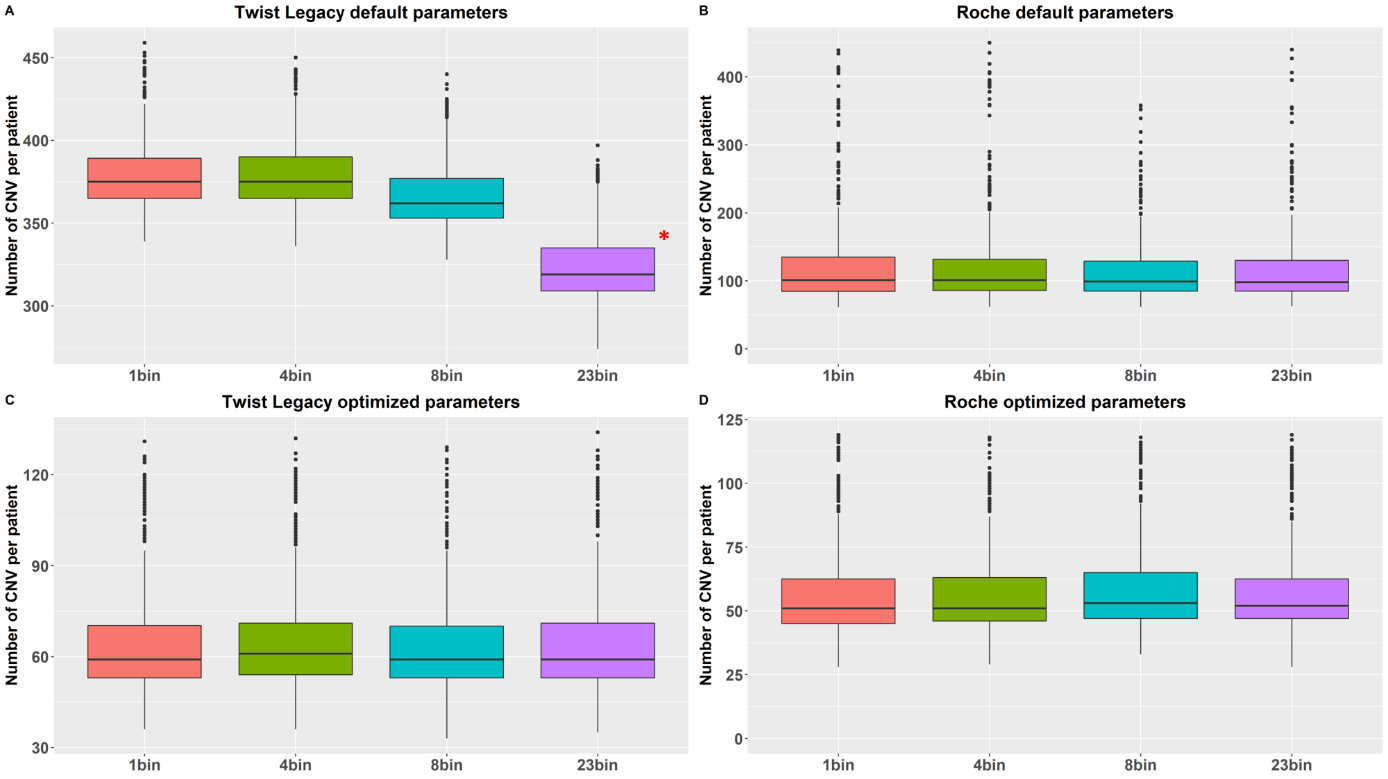

**Supplementary Figure 3**. Distribution of the CNV number per patient according to the parameter set, the kit used and the number of bins.

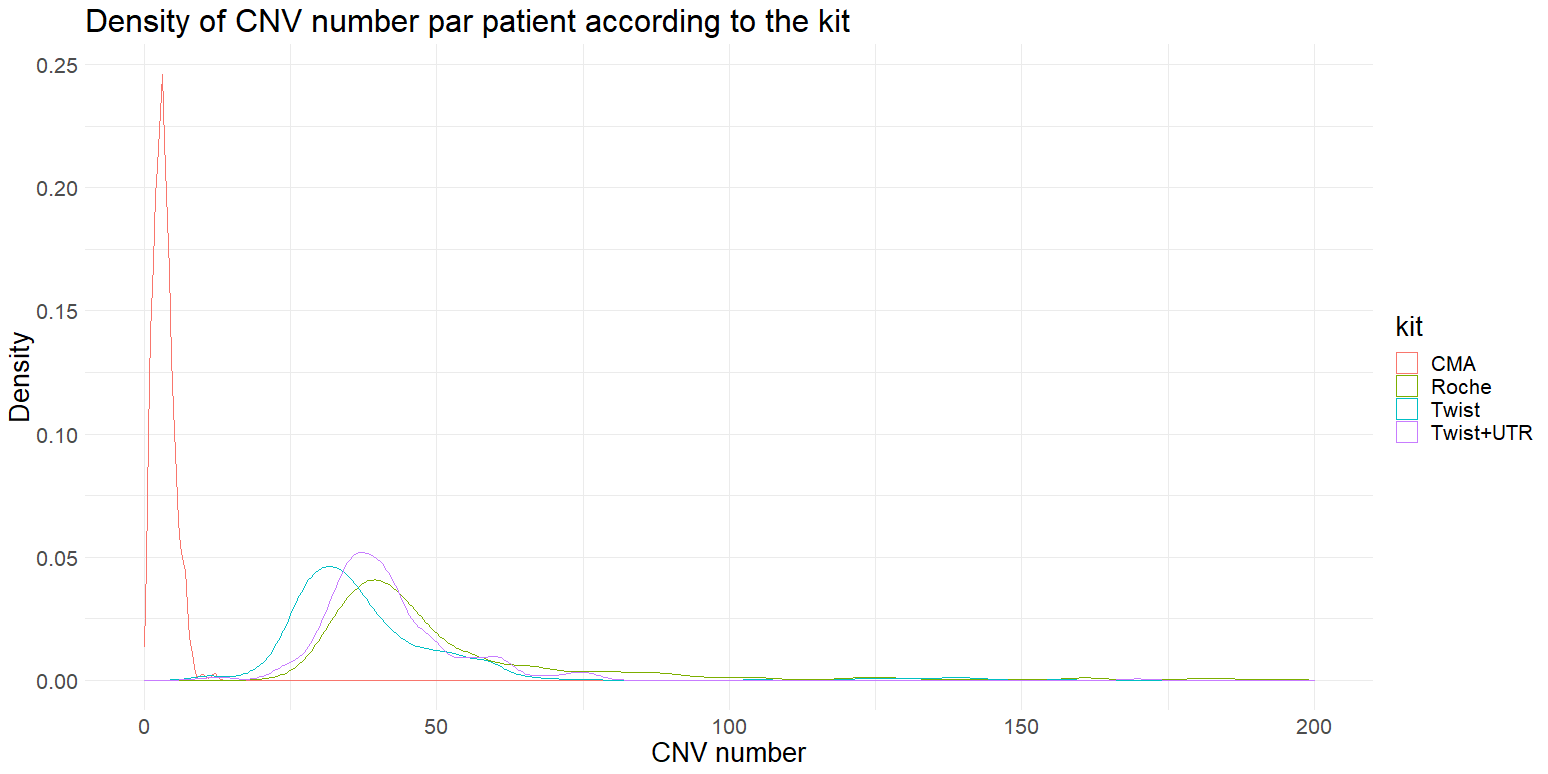

**Supplementary Figure 4.** Density of CNV counts per patient according to the kit / CNV model. *CMA (n=300), Roche (n=511), Twist (n=1154), Twist UTR (n=383).*

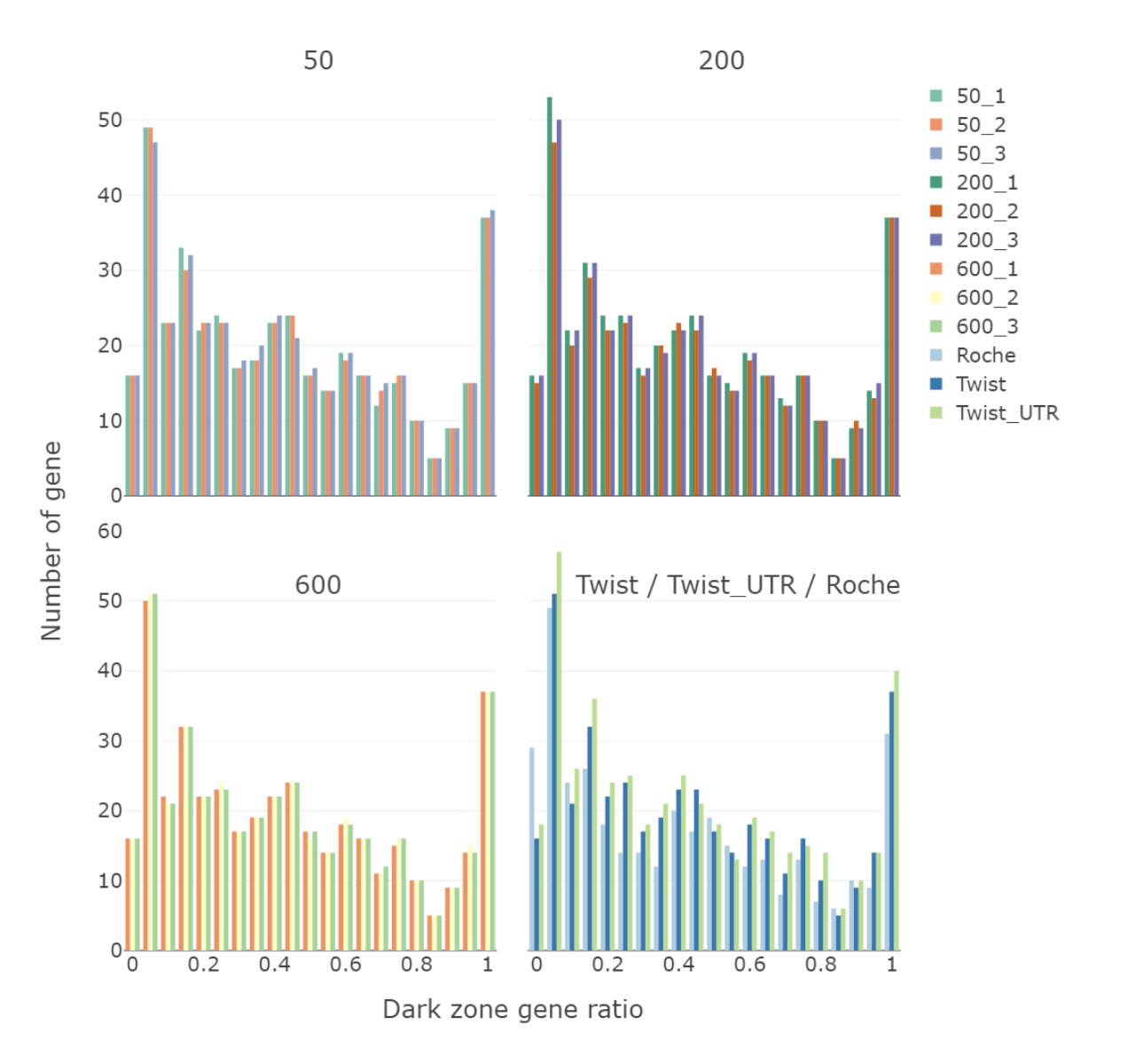

**Supplementary Figure 5**. Proportion of morbid genes coding sequence in CNV calling pipeline models black area of. Each color represents the number and proportion of genes in the black zone per model. Results shown from populated models of 50, 200, and 600 samples sequenced with the Twist Human Core exome kit, as well as results from Twist (n =1154), Roche (n=511), and Twist+UTR (n=383) models. 1 : 100% of the coding sequence is in the black zone.

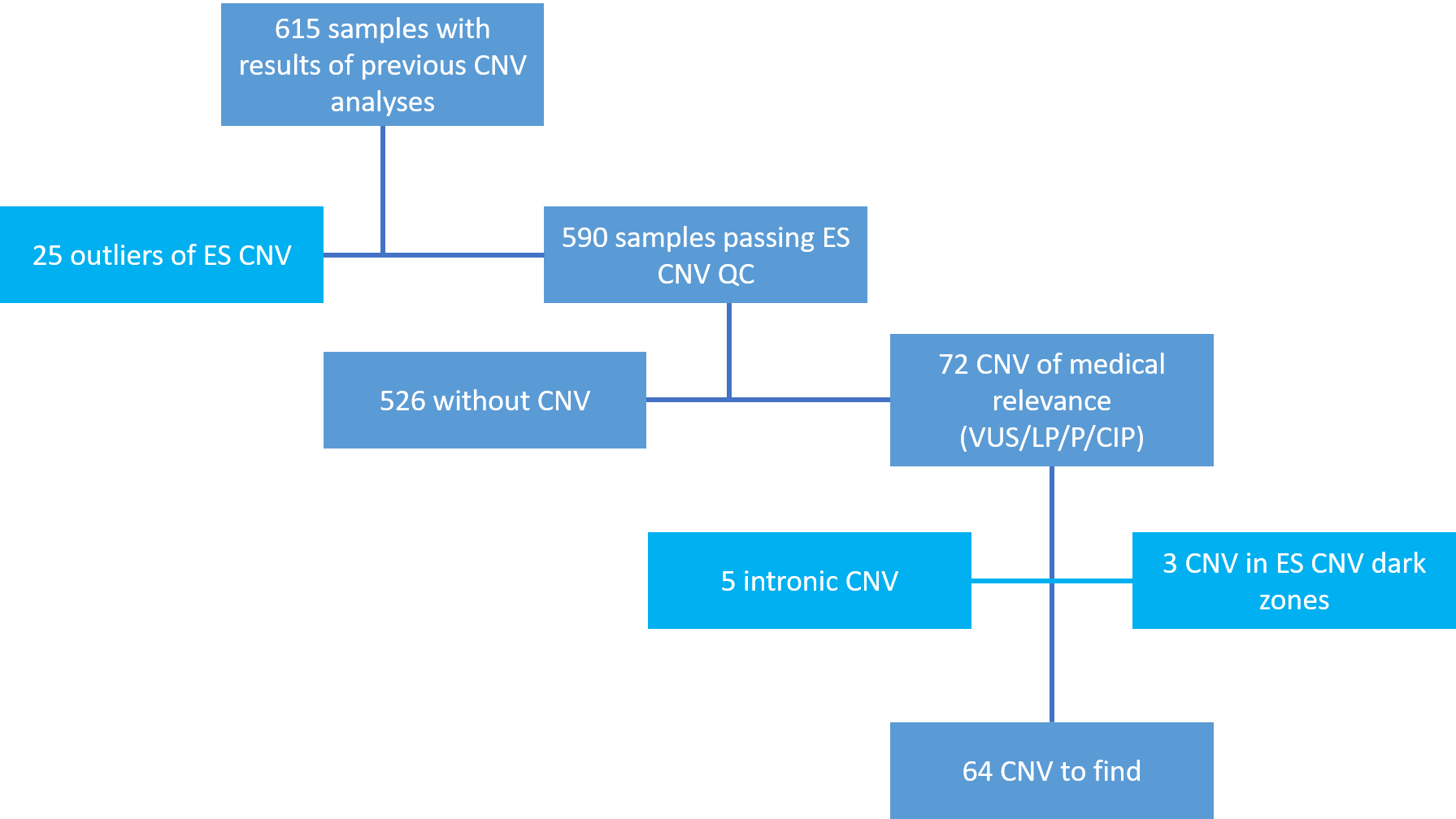

**Supplementary Figure 6**. Methodology for determining the validation cohort CNVs from pre-existing CMA data. Light blue : CNVs eliminated from the validation cohort. VUS : Variant of Uncertain Significance, LP : Likely Pathogenic, P : Pathogenic, CIP : CNV with incomplete penetrance.

|  | RD | MCA/IDD | Total |
| --- | --- | --- | --- |
| Roche | 59 | 388 | 447 |
| Twist UTR | 249 | 739 | 988 |
| Twist | 323 | 660 | 983 |
| Total | 631 | 1787 | 2418 |

**Supplementary Table 4. Details of the two subgroups of patients sequenced in this study.** RD : Renal Diseases, MCA : Multiple Congenital Anomaly, IDD : Intellectual Disability Disorder.
